# Cross-sectional Ct distributions from qPCR tests can provide an early warning signal for the spread of COVID-19 in communities

**DOI:** 10.1101/2023.01.12.23284489

**Authors:** Mahfuza Sharmin, Mani Manivannan, David Woo, Océane Sorel, Jared Auclair, Manoj Gandhi, Imran Mujawar

**Affiliations:** Thermo Fisher Scientific, 180 Oyster Point Blvd, South San Francisco, CA 94080, USA; Northeastern University, 147 S. Bedford St, Burlington, MA 01803, USA

## Abstract

**Background:** SARS-CoV-2 PCR testing data has been widely used for COVID-19 surveillance. Existing COVID-19 forecasting models mainly rely on case counts, even though the binary PCR results provide a limited picture of the pandemic trajectory. Most forecasting models have failed to accurately predict the COVID-19 waves before they occur. Recently a model utilizing cross-sectional population cycle threshold (Ct) values obtained from PCR tests (Ct-based model) was developed to overcome the limitations of using only binary PCR results. In this study, we aimed to improve on COVID-19 forecasting models using features derived from the Ct-based model, to detect epidemic waves earlier than case-based trajectories.

**Methods:** PCR data was collected weekly at Northeastern University (NU) between August 2020 and January 2022. The NU campus epidemic trajectories were generated from the campus incidence rates. In addition, epidemic trajectories were generated for Suffolk County, where NU is located, based on publicly available case-counts. A novel forecasting approach was developed by enhancing a recent deep learning model with Ct-based features, along with the model’s default features. For this, cross-sectional Ct values from PCR data were used to generate Ct-based epidemic trajectories, including effective reproductive rate (Rt) and incidence. The improvement in forecasting performance was compared using absolute errors and residual squared errors with respect to actual observed cases at the 7-day and 14-day forecasting horizons. The model was also tested prospectively over the period January 2022 to April 2022.

**Results:** Rt estimates from the Ct-based model preceded NU campus and Suffolk County cases by 12 and 14 days respectively, with a three-way synched Spearman correlation of 0.57. Enhancing the forecasting models with Ct-based information significantly decreased absolute error and residual squared error compared to the original model without Ct features (p-value <0.001 for both 7 and 14-days forecasting horizons).

**Conclusion:** Ct-based epidemic trajectories can herald an earlier signal for impending epidemic waves in the community and forecast transmission peaks. Moreover, COVID-19 forecasting models can be enhanced using these Ct features to improve their forecasting accuracy.

**Policy implications:** We make the case that public health agencies should publish Ct values along with the binary positive/negative PCR results. Early and accurate forecasting of epidemic waves can inform public health policies and countermeasures which can mitigate spread.

## INTRODUCTION

The COVID-19 pandemic created an unprecedented situation where large-scale PCR testing has been performed to identify cases who were suspected to be infected with the virus. The results from such widespread testing have also helped better understand the epidemiological aspects of this disease and adapt public health interventions to reduce the spread of the virus. Current approaches for outbreak surveillance rely mainly on SARS-CoV-2 PCR testing results to estimate case counts and test positivity rates^1^.While real-time or near real-time monitoring at the local and national levels can help detect changes in patterns of the pandemic and inform public health decisions, this approach, however, does not allow public health agencies sufficient lead time to prepare for upcoming surges. Public health agencies need to gain access to data predicting the evolution of the pandemic as early as possible in order to plan for surges and adapt healthcare capacities accordingly^2^.

Concerted efforts from academia, industry, and government have been made to develop predictions at different forecasting horizons. Notably, these efforts has led to development of the COVID-19 Forecast Hub, which provides up-to-date data of COVID-19 cases, deaths and hospitalizations in the US, in coordination with the Centers for Disease Control and Prevention (CDC) ^3^. Most of these forecasting initiatives use various compartmental models^4,5^ a modelling technique where populations are clustered into various compartments such as susceptible, infectious, and recovered, with flow patterns being simulated between them. However, almost all of these compartmental models rely mainly on the binary (i.e., positive/negative) test results and provide a limited picture of the pandemic trajectory^6^.Case-series trajectories, based on incidence rates alone, can exhibit biases due to variations in testing demand and supply, population sampling and reporting delay^7,8^. Most forecasting models currently available on the Forecast Hub have struggled to consistently predict COVID-19 waves before they occur^3^. The ensemble model which combines the top performing models, developed by experts in the field, was unable to detect any of the variant waves in the US. This model has shown “low reliability” in predicting COVID-19 waves and is no longer used ^9^.These findings highlight the tremendous challenge of predicting COVID-19 surges with forecasting models using the binary test outcomes.

Recently, Hay et al. ^6^ have shown that using population cross-sectional viral load distributions, measured by PCR cycle thresholds (Ct), can overcome common biases in epidemic trajectory estimates that are seen when using case series data alone. Additional studies have corroborated this observation and showed that the models with Ct information correlated to case counts^10^ and can be used to better predict epidemic growth rates^11,12^. In this study, we further corroborate that Ct-based model-derived epidemic trajectories can detect epidemic waves earlier than case-based trajectories, and that COVID-19 forecasting models can be enhanced using these Ct features to improve their forecasting accuracy.

## METHODS

### qPCR data preparation

During the first couple years of the pandemic, the Life Science Testing Center (LTSC) conducted routine PCR screening of students, faculty and staff at Northeastern University using the TaqPath™ COVID-19 Combo Kit. In this study, PCR data were retrospectively analyzed from samples collected between August 2020 and January 2022, and prospectively up to April 2022. Deidentified .*eds* files from the PCR runs were analyzed to obtain the Ct values from all screened samples during the defined period. According to the TaqPath™ COVID-19 Combo Kit Instructions for Use, a sample was considered positive for SARS-CoV-2 if two or more of the three gene targets (N, S and ORF1ab) were detected (Ct≤37). Samples where only one gene target was detected (Ct≤37) were considered as inconclusive^13^.

### Incidence for NU campus and Suffolk County case-series

Daily campus COVID case-count was determined through counting the number of unique positive samples (adjusting for re-testing) per day. Since the data was de-identified, these campus case-counts were not adjusted for different samples taken from the same individual or for re-infection. Incidence per 100,000 population was calculated by dividing cases per day or week by the total campus population as of 2019^14^. COVID-19 cases in Suffolk county, where the university campus is located, were obtained from a publicly available database^15^. Incidence per 100,000 population was calculated by dividing the county positive cases by the county’s total population based on census numbers as of April 2020^16^.

### Ct-based model to determine inferred incidence estimates

Incidence curves were created using the Ct values from the PCR screening tests performed on campus according to the methodology developed by Hay et. al. ^6^. Only the N-gene Ct values from the TaqPath COVID-19 Combo Kit were selected to be included in the Ct-based model. Based on their run dates, the samples were considered as weekly occurrences. We used a Gaussian process model from the *virosolver* R-package^17^ the weekly Ct-distributions to infer the incidence rates and followed the recommended parameters for the Gaussian process model. In the initial phase of the study, we retrospectively analyzed the epidemic trajectories from the Ct-based model for early detection of incidence trends of the campus and county from the samples collected between August 2020 to January 2022. In the next phase, we prospectively monitored the ability of the Ct-based model for early indicators of uptick in incidence trends in NU and Suffolk county on a weekly basis from January 2022 until April 2022.

### Estimating time varying reproduction number (Rt)

The three incidence trajectories: a) Ct-based model derived estimates, b) campus positive samples, and c) county case-series, were smoothed by a 14-day moving average. The effective reproduction number (Rt) was derived from these three incidence trajectories, using the R-package *EpiEstim*^18^. The *EpiEstim* package estimates the effective reproduction number of an epidemic, given the incidence time series and the serial interval distribution. We used a parametric serial intervention method with a mean serial interval of 6.14 and standard deviation of 3.96 following recommended parameters, and a 14-day sliding window. The data flow and processing steps are depicted in Fig. 1a.

**Figure 1.**
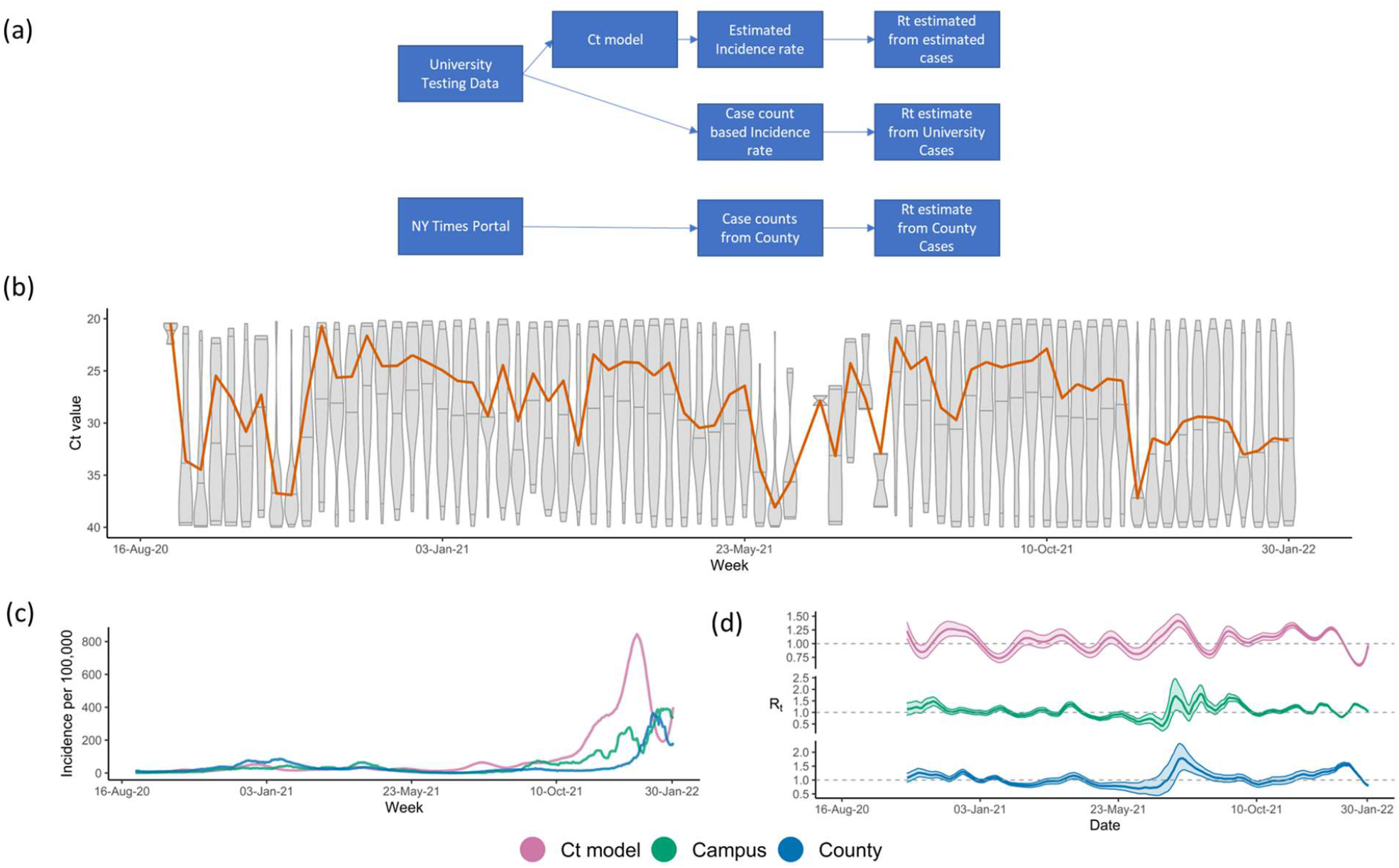
**(a)** Study design showing data flow and processing; **(b)** Weekly Ct value distribution represented by each boxplot. The orange line represents the rolling median of the weekly Ct distribution; **(c)** Incidence rates predicted from Ct-based model (magenta), campus case counts (green), and Suffolk County case counts (blue); **(d)** Rt estimates derived from Ct model inferred incidences (magenta), or campus (green) and Suffolk County case counts (blue).

### Lagged Correlation Analysis between Ct-based model Rt curves, Campus case-based Rt curves, and County case-based Rt curves

Pairwise lagged correlation was assessed to estimate the Spearman Correlation Coefficients and the lag or lead times between the 3 Rt curves: Ct-based model, campus case-based, and county case-based. Transmission peaks were identified when the Rt estimates were greater than 1. Transmission peaks in the Ct-based model Rt curves were mapped to subsequent transmission peaks detected in the campus and county Rt curves. The number of days between the Ct-based model peak and the corresponding presumptive subsequent peak in the campus and county curves was calculated. The median time to campus and county peak from a Ct-based model Rt peak was then estimated.

### Neural relational autoregressive model (β-AR) and Ct-enhanced β-AR (Ct-β-AR)

β-AR is a case count forecasting model that was published by Facebook/Meta^19^.The β-AR model requires two types of inputs: a) time dependent data including symptom survey, mobility, doctor visits, total PCR tests per state, vaccination and masking policy, and weather, which are modeled by the recurrent neural network (RNN) part of the overall model and b) confirmed COVID-19 cases variable is the only time independent input, which feeds into the autoregressive (AR) part of the model. We enhanced the β-AR model using features derived directly from PCR Ct values and from the Ct-based model above. Each feature was measured weekly in accordance with the β-AR model measurement frequency.

### Determining accuracy of Ct-β-AR to real incidence rates using RMSE and AE

We assessed performance of Ct-β-AR and β-AR from October 2020 to December 2021, by comparing the predictions at the 7- and 14-day forecast horizons to real incidence rates. To measure the performance, we calculated the root mean square error (RMSE) and mean absolute error (AE) by the following formula:

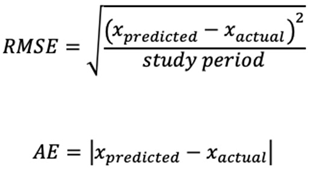

where x_actual_ was the actual case count on a particular day and x_predicted_ was the predicted case count for the same day. The difference between Ct-β-AR and β-AR models for their RMSE and AE values were statistically assessed using the paired Wilcoxon signed rank sum test.

## RESULTS

### Data cohort with PCR results description

A total of 1,608,562 valid samples, collected between August 2020 and April 2022, were included in the analysis. Among those samples, 47,194 samples tested positive for SARS-CoV-2, and 17,677 samples were identified as inconclusive. Over the study period, the median N-gene detectable Ct value was around 28 with a minimum of 5 and a maximum of 40 and a skewness of -5.69. During the *wild-type* (initial Wuhan) (Aug 2020 – Jan 2021) N=154; *Alpha* (Jan 2021 – Jun 2021) N=7,216; *Delta* (Jul 2021 – Dec 2021) N=11,790; and *BA*.*1 Omicron* (Jan 2022 – Mar 2022) N=21,715 surges, samples tested positive with an approximate median Ct=28 across waves and varying skewness of -9.07, -7.38, -5.13, -3.22, respectively. The Ct distributions over time are shown in Fig. 1b.

### Ct-based and Case-count derived epidemic metrics were highly comparable

The predicted incidence rates, derived from either Ct-based model, Campus or Suffolk County cases counts (Fig. 1c) were very similar in terms of epidemic trends throughout the duration (Aug 2020 to Jan 2022). Upsurges in incidence were observed around the same periods for the 3 incidence trajectories, including when the *wild-type* variant was dominant as well as during the *Alpha, Delta*, and *Omicron* variant waves. The trajectories also demonstrated similar intervening troughs between the observed waves. The incidence waves were also relatively similar in size, apart from the first Omicron wave, where the Ct-based incidence wave was much larger than the other two trajectories. The three incidence rate patterns were also consistent with publicly available data for Massachusetts state^20^ showing that the predicted values were in agreement with the local epidemic incidence rates. The effective reproduction number (Rt) estimates for the 3 corresponding incidence trajectories were also very similar throughout the period analyzed (Fig. 1d). We observed that major Rt peaks noted at the beginning of a variant wave generally matched across the 3 trajectories, especially between the Ct-based and county Rt trajectories. The synched Spearman Correlation Coefficient between the three Rt curves (Ct-based vs Campus case count vs County case count) was 0.57 demonstrating that the Ct-based epidemic trajectory was highly comparable to the case-count derived epidemic trajectories (Campus and County), considering the duration analyzed (18 months) and a 3-way synched correlation.

### Ct-based model predictions preceded case counts-derived estimates

Once we established that the Ct-based model was able to accurately mirror the epidemic trends, we proceeded to determine if the Ct-based model was able to portend impending surges over case-count derived models, and if so, how early could our model predict. Lagged correlation analysis of the Rt estimated curves showed that the Ct-based model had a 12-day lead time as compared to the Campus case count estimates and a 14-day lead time as compared to County case count estimates, with lagged Spearman correlation coefficients of 0.38 and 0.59 respectively. Analysis of the Rt trend peaks showed that Ct-based model peaks preceded the corresponding case counts-derived Rt peaks by a median lead time of 11 days for, both, the county (Fig. 2. a, b) and campus estimates (Fig. 2c, d). Interestingly, prospective monitoring of the Rt trends showed that the Ct-based model derived Rt trajectory was able to detect an upcoming peak around week 4 of March 2022 which was not yet uncovered in, both, the case counts based Rt trajectories (Fig. 3a). Upon further monitoring, it was discovered that the Ct-based model predicted peak was, indeed, followed by a peak in actual Campus and County case-counts after 2 weeks (week 2 of April 2022) which was due to the *Omicron* surge (BA.2) (Fig. 3b).

**Figure 2.**
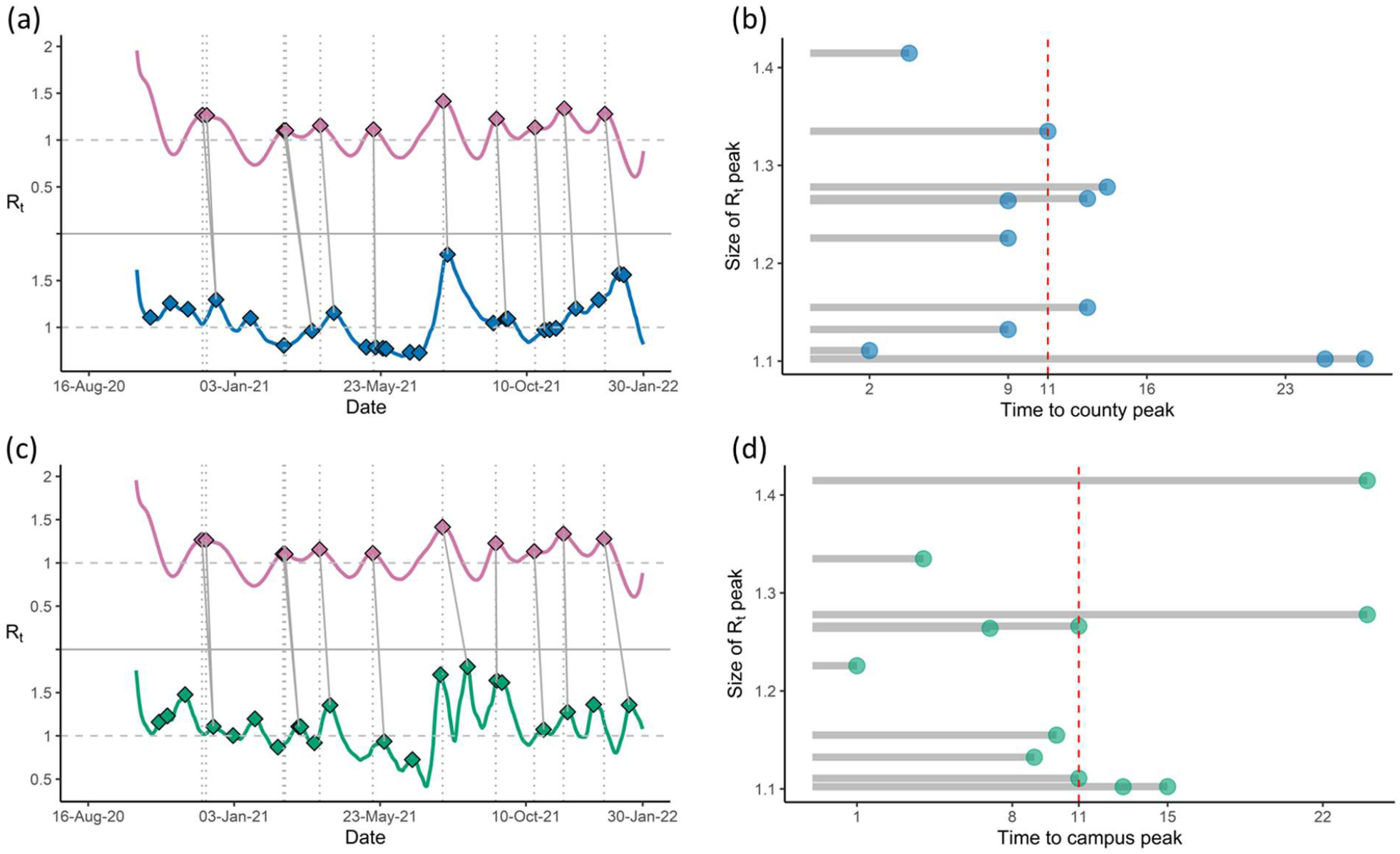
**(a)** R_t_ trends showing peaks for Ct-based model derived estimates (magenta), with subsequent peaks in R_t_ trends derived from county cases (blue); **(b)** Lead times (in days) between Ct model-derived R_t_ peaks and county R_t_ peaks, with median lead time (red dashed line). **(c)** R_t_ trends showing peaks for Ct model-derived estimates (magenta), with subsequent peaks in R_t_ trends derived from positive tests on campus (green); **(d)** Lead times (in days) between Ct-based model derived R_t_ peaks and campus R_t_ peaks, with median lead time (red dashed line).

**Figure 3.**
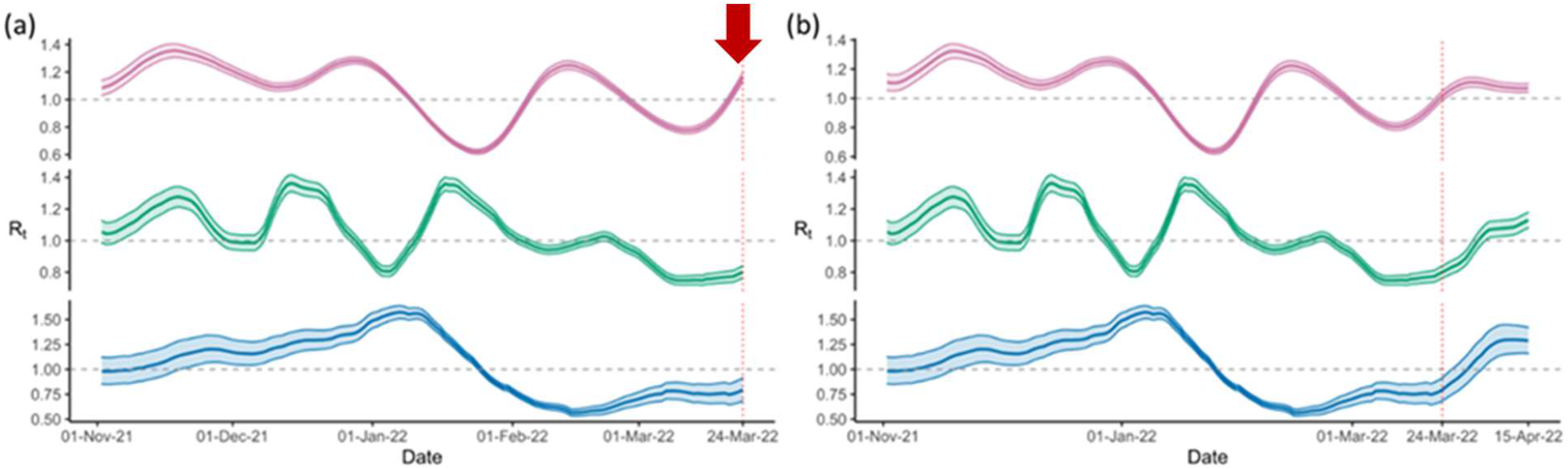
Prospective monitoring of the Rt trajectories showed **(a)** R_t_ trends with an early uptick (red arrow) in Ct model-derived estimates (magenta) around 24 March 2022, while the other two trajectories, including R_t_ trends derived from county cases (blue), and positive tests on campus (green) are flat; **(b)** Later upticks noted in R_t_ trends derived from county cases (blue), and positive tests on campus around 15 April 2022.

### Ct-β-AR model significantly improved forecasting estimates

Over the time duration analyzed (October 2020 to December 2021), the Ct-β-AR model provided statistically significant lower AE at both forecasting horizons; 7 days (*p*-value <0.001) (Fig. 4a, b) and 14 days (*p*-value <0.001) (Fig. 4c, d). In addition, the mean AE values comparing Ct-β-AR vs. β-AR models were 273.4 vs. 322.8 and 679.6 vs. 901.1 for the 7 and 14-day forecasting horizons, respectively. Similarly, the RMSE was also significantly lower (*p*-value <0.001) comparing Ct-β-AR vs. β-AR with RMSE values of 366.3 vs. 406.6 and 828.8 vs. 1045.9 for the 7 and 14-day forecasting horizons, respectively. Regardless of which analytical method was used, RMSE or AE, enhancing the model with Ct features reduced the error to the real incidence value and provided more accurate forecasting than the β-AR model.

**Figure 4:**
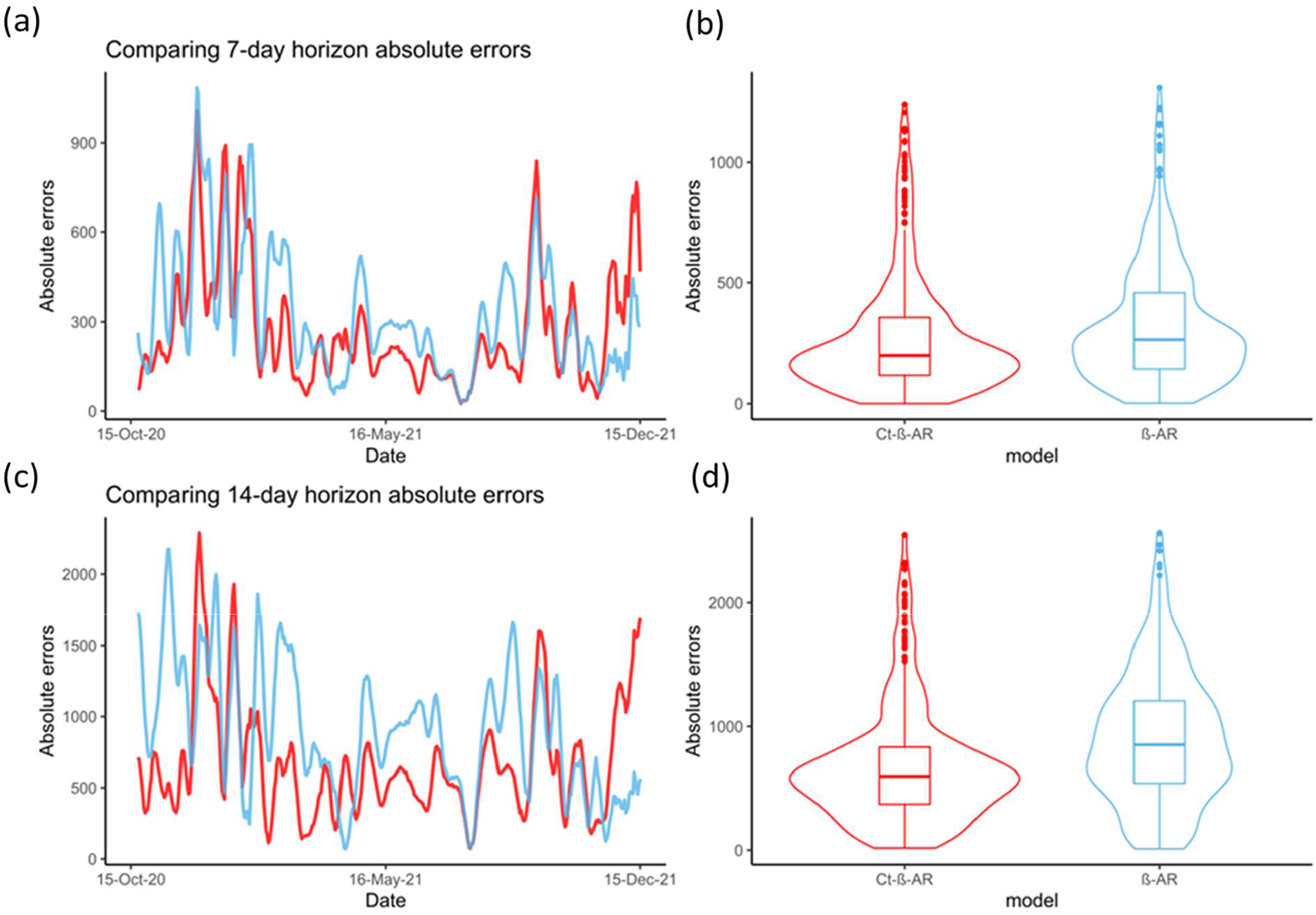
(a) Comparing trends of 7-day horizon absolute errors for Ct-β-AR (red) and β-AR (blue) models; (b) Comparing distributions of 7-day horizon absolute errors for Ct-β-AR (red) and β-AR (blue) models; (c) Comparing trends of 14-day horizon absolute errors for Ct-β-AR (red) and β-AR (blue) models; (d) Comparing distributions of 14-day horizon absolute errors for Ct-β-AR (red) and β-AR (blue) models;

## DISCUSSION

In this study, we demonstrate that Ct-derived information from routine PCR testing can detect epidemic waves earlier than case-based trajectories, and by doing so, provides an earlier indication of upcoming COVID-19 waves in the community. We took the β-AR forecasting model which uses deep learning and autoregression to produce weekly forecasts up to a 4-week horizon and enhanced it with Ct features from PCR test results. The model enhanced by Ct-derived features provided more accurate forecast (lower RMSE/AE) than the conventional approach.

Our observations are consistent with three previous studies and further corroborate and highlight the value of Ct values in epidemic forecasting^10-12^. The study conducted in Madagascar showed that epidemic growth curves integrating Ct-derived features slightly preceded incidence data-derived epidemic growth curves^12^. Likewise, the study conducted in Belgium that correlated daily/weekly Ct value distributions and case counts showed that epidemic trajectories incorporating Ct values preceded those derived by case counts by 17 days^11^. Lastly, the Hong Kong study, observed 7 days of leading Rt waves from the log-regression model derived from the Ct statistics as compared to the Rt waves derived from classic case counts^10^. Our study adds to this body of evidence that highlights the importance of Ct values for epidemic trajectories and how they could be leveraged to predict surges in a timely manner.

Some of the more widely used forecasting models use features such as population mobility data, international flight data, government restrictions, non-pharmaceutical interventions, vaccination, and seasonality, as well as COVID-19 symptoms survey data^19,21-24^. With the exception of COVID-19 symptom data, most of these features such as international flight data, government restrictions etc. are not grounded in the inherent biology of the infection. There are other forecasting methods that have been developed that take some of the biological characteristics of the virus into account; for example, wastewater surveillance^25^ that measures concentrations of the virus in wastewater to infer community-level disease dynamics, or self-reported COVID-19 symptoms^26^, or detecting pre-symptomatic cases using digital wearable devices^27,28^, or a combination of self-reporting and digital wearable devices^29^. However, as compared to these models, Ct values would still be the most readily available since they are automatically generated with most routine COVID-19 RT-PCR tests.

While our Ct-enhanced model showed better forecasting than the case-count derived forecasting approach, our study is not without limitations. Firstly, our data were obtained from a single PCR assay. However, Hay et. al demonstrated that the using different PCR methods does not influence the ability the Ct-based model to accurately estimate the epidemic trajectory. Secondly, currently used forecasting models can make predictions at different geographical levels such as at the national, state^23^ [and/or the county level^30-32^ while we assessed our model at a smaller geographical scale, at the community and the county level. Thirdly, we enhanced only one forecasting model, the β-AR model, to assess the usefulness of incorporating Ct values. There are other publicly available forecasting models employing a compartmental model methodology ^21,23,24,33-35 36 37,38^, machine learning techniques^39 30,40^, deep learning approaches^41-43, 44, 44,45^, ensemble methods ^9,46,47^, statistical methods^48-51^, hybrid approaches^52,53 54 55-57^ that could be enhanced to validate our approach.

More importantly, what this study, along with others, has shown is the potential of utilizing PCR Ct values for surveillance of other respiratory infectious diseases such as seasonal influenza or its broader use in the field of other infectious diseases. The value of PCR Ct data has often been overlooked for public health surveillance due to lack of awareness and/or due to the lack of data availability. There is now growing evidence that Ct-derived forecasting methods are an effective method for sentinel surveillance and early identification of surges. Many groups have advocated for public reporting of Ct information for surveillance, especially in countries with limited public health surveillance resources with routine testing laboratories even acting as “epidemic sensors” for the geographical regions they serve^11,12^. Given the compelling evidence for the use of Ct values in disease monitoring and forecasting, we strongly urge public health agencies to make PCR Ct values publicly available along with the binary test results.

## Data Availability

All data produced in the present work are contained in the manuscript

## Notes

### Competing Interest Statement

The authors declare the following competing interests: MS, MM, DW, OS, MG and IM are employees of Thermo Fisher Scientific a corporation that funded the study.

### Funding Statement

This study was funded by Thermo Fisher Scientific and Northeastern University's Life Sciences Testing Center

### Author Declarations

All samples were deidentified for study purposes in accordance with IRB approval by the Northeastern 223 University Office of Human Subject Research Protection (HSRP).

